# Evidence-Based Practice Competencies among Nutrition Professionals and Students: A Systematic Review

**DOI:** 10.1101/2023.08.03.23293580

**Authors:** Nirjhar R. Ghosh, Zahra Esmaeilinezhad, Joanna Zając, Rebecca A. Creasy, Saundra G. Lorenz, Molly Crews, Karen M. Beathard, Malgorzata M. Bala, Kevin C. Klatt, Bradley C. Johnston

## Abstract

**Background:** Evidence-based practice (EBP) promotes shared decision-making between clinicians and patients and has been widely adopted by various health professions including nutrition & dietetics, medicine and nursing.

**Objective:** To determine evidence-based practice (EBP) competencies among nutrition professionals and students reported in the literature.

**Design:** Systematic review.

**Data sources:** Medline, Embase, CINAHL, ERIC, CENTRAL, ProQuest Dissertations and Theses Global, BIOSIS Citation Index, and ClinicalTrials.gov up to March 2023.

**Eligibility criteria for study selection:** Eligible primary studies had to objectively or subjectively document the assessment of at least one of six predefined core EBP competencies, including formulating structured clinical questions, searching the literature for best evidence, and assessing studies for methodological quality, magnitude (size) of effects, certainty of evidence for effects, and determining the clinical applicability of study results based on patient values and preferences.

**Data extraction and synthesis:** Two reviewers independently screened articles and extracted data, including the reporting quality for eligible studies. Results were not amenable to meta-analysis and were thus summarized for each EBP competency.

**Results:** We identified 12 eligible cross-sectional survey studies, comprised of 1065 participants, primarily registered dietitians, across six countries, with the majority assessed in the United States (n=470). The reporting quality of the survey studies was poor overall, with 43% of items not reported and 22% of items partially reported. Only one study (8%) explicitly used an objective questionnaire to assess EBP competencies. The proportion of studies reporting on each competency were: 17% on the formulation of clinical questions, 83% on searching the literature, 75% on methodological quality or critical appraisal, 58% on interpreting statistical results, and 75% on applying study results. In general, the six competencies were incompletely defined or reported (e.g., it was unclear what ‘applicability’ and ‘critical appraisal’ referred to, and what study designs were appraised by the participants). Two core competencies, the magnitude (size) of effects and the certainty of evidence for effects, were not assessed.

**Conclusions:** Among 12 included articles the overall quality of study reports was poor, and when EBP competencies were reported they were predominantly self-perceived assessments as opposed to objective assessments. No studies reported on competencies in assessing magnitude of effect or certainty of evidence, skills that are essential for optimizing clinical nutrition decision-making.

**Systematic review registration:** PROSPERO CRD42022311916.

## INTRODUCTION

Evidence-based practice (EBP) promotes shared decision-making between clinicians and patients based on three foundational principles: (i) the use of best available evidence, (ii) clinical or real-world experience, and (iii) the consideration of patients’ values and preferences based on their unique circumstances[1,2]. Having originated from the concept of evidence-based medicine (EBM) first described by Guyatt in 1991[3,4], EBP has been widely adopted by various health professions[5–9]. Based on the Users’ Guides to the Medical Literature[10], EBP core competencies can be described as (i) formulating a structured clinical question, (ii) finding the best available research evidence, (iii) assessing the methodological quality or risk of bias (RoB) of available evidence, iv) assessing the study results (i.e., magnitude (size) and precision of effects) for all desirable (benefits) and undesirable (harms) outcomes, and (v) applying results to clinical care based on the generalizability of the evidence to one’s patients, including the patients’ values and preferences based on the evidence for potential benefits, harms and the burdens of an intervention. Here, values refer to the relative worth, merit or importance of health outcomes to the patients (e.g., mortality vs non-fatal stroke vs blood pressure), and based on the outcomes patients most value, preferences refer to patients’ preferred treatment choices after the best available evidence for alternative management strategies is shared with them[11].

According to the International Confederation of Dietetic Association’s definition, “*Evidence-based dietetics practice involves the process of asking questions, systematically finding research evidence, and assessing its validity, applicability, and importance to nutrition and dietetics practice decisions; and applying relevant evidence in the context of the practice situation, including professional expertise and the values and circumstances of patients/clients, customers, individuals, groups, or populations to achieve positive outcomes*.”[12] Similarly, the U.S. Accreditation Council for Education in Nutrition and Dietetics states in their 2022 Accreditation Standards that the curricula of Didactic Programs in Dietetics and Dietetic Internship should prepare dietetic students and interns to locate, interpret, evaluate, and use peer-reviewed nutrition literature to make evidence-based practice decisions[13]. Comparably, the ‘Partnership for Dietetic Education and Practice’ in Canada, the ‘National Competency Standards for Dietitians’ in Australia and the British Dietetic Association state in their accreditation standards or curriculum framework that the dietetic programs should equip dietitians with the ability to employ or demonstrate evidence-based approaches to dietetic practices[14–16]. Given the global EBP mandate set forth by leading dietetic associations[13–16] and the precedent of assessing EBP competencies in other health professionals[17], it is timely to evaluate the knowledge, skills, attitudes, and behaviors of nutrition professionals and students regarding EBP competencies that are believed to facilitate informed decision-making between patients and clinicians. Therefore, we systematically reviewed and documented the totality of published evidence assessing different EBP competencies among nutrition professionals and students.

## METHODS

### Search methods for identification of primary studies

We searched five electronic databases: Medline, Embase, CINAHL, ERIC and CENTRAL, from inception to March 2023. In addition, we searched the gray literature using ProQuest Dissertations and Theses Global, BIOSIS Citation Index, and ClinicalTrials.gov up to March 2023. Reference lists of included studies were searched to help ensure all eligible studies were identified. We did not restrict our search based on language of publication or publication status. See **Appendix A** for detailed Medline search strategies. Full search strategies are available on request. We followed Preferred Reporting Items for Systematic Reviews and Meta-Analyses (PRISMA) 2020 statement[18] and Synthesis without meta-analysis (SWiM)[19] to report our review and the protocol was registered in PROSPERO (CRD42022311916)[20].

### Definition of variables

Regarding analyzing our data, we used the term ‘outcomes’ to refer to the broader knowledge, skills, attitudes, and behaviors relevant to EBP competencies. We defined *knowledge* as the depth of learner’s awareness and understanding of EBP concepts; *skills* as the ability to apply knowledge and perform EBP steps in a practical setting; *attitudes* (also *perceptions, confidence* and *willingness*) indicated how individuals perceived the importance of EBP, including their willingness to apply EBP principles; and *behaviors* referred to one’s real-life execution of EBP steps[21,22]. We used the term ‘competency’ to indicate the specific domains of EBP. That is, one needs to have knowledge, skills, attitudes, and behaviors in specific domains (e.g., formulating answerable clinical questions, assessing various study designs for methodological quality) to apply EBP effectively.

We elected to use the five EBP competencies based on the Users’ Guides to the Medical Literature[10], while adding one additional competency based on a recent consensus statement on core EBP competencies for health professionals[23]. This sixth competency addresses interpreting the certainty of evidence for outcomes of benefit and harm based on study results, ideally based on up-to-date high-quality systematic reviews with meta-analysis and/or guidelines based on such reviews[22]. See summary of six competencies in **Table 1**.

**Table 1.**
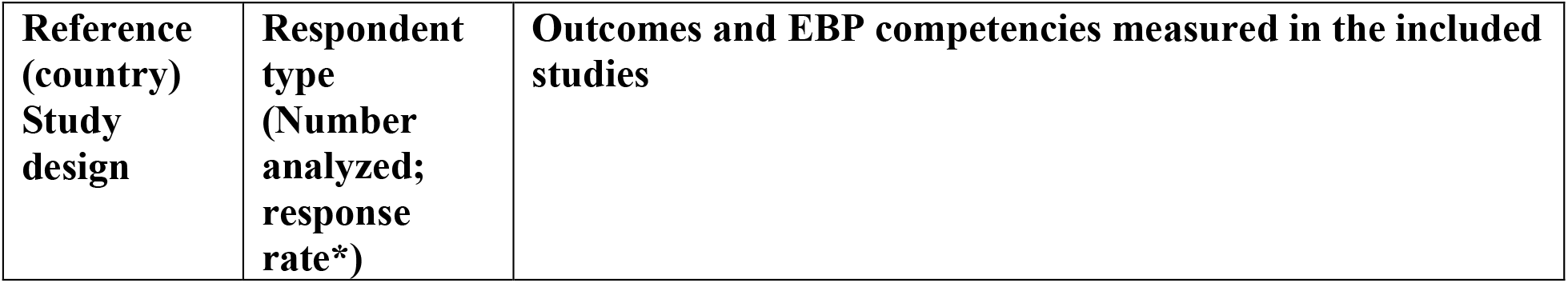
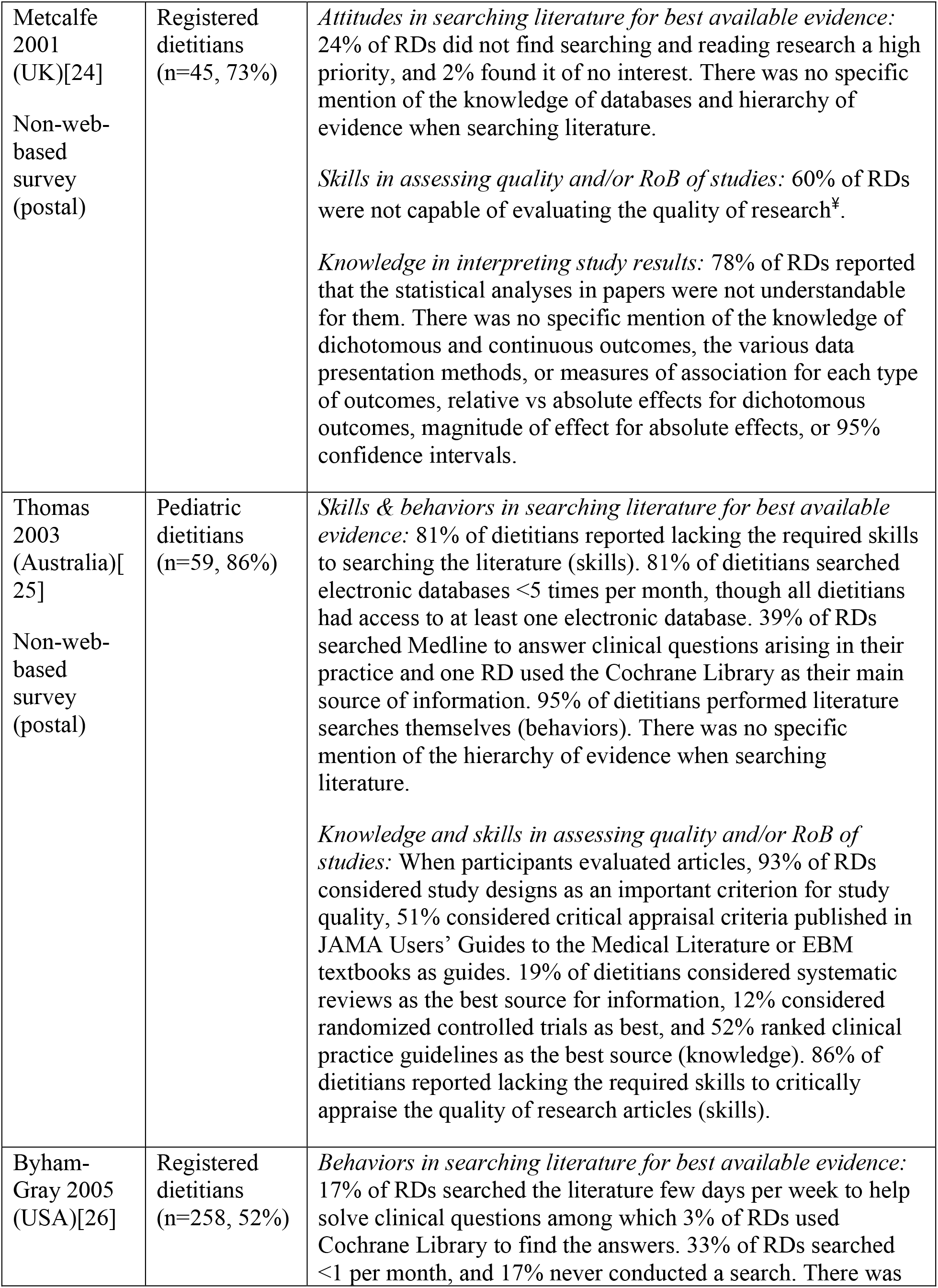
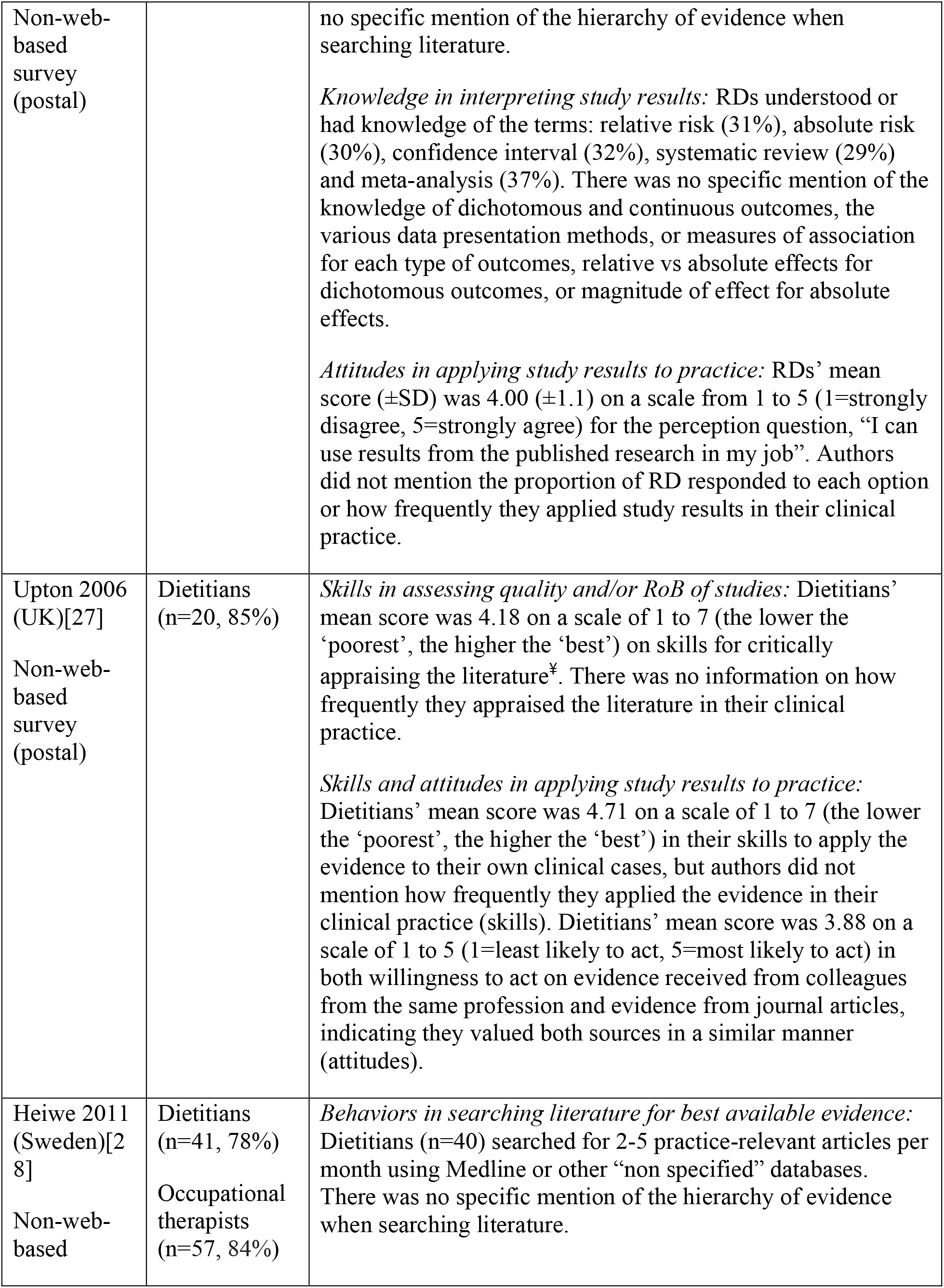
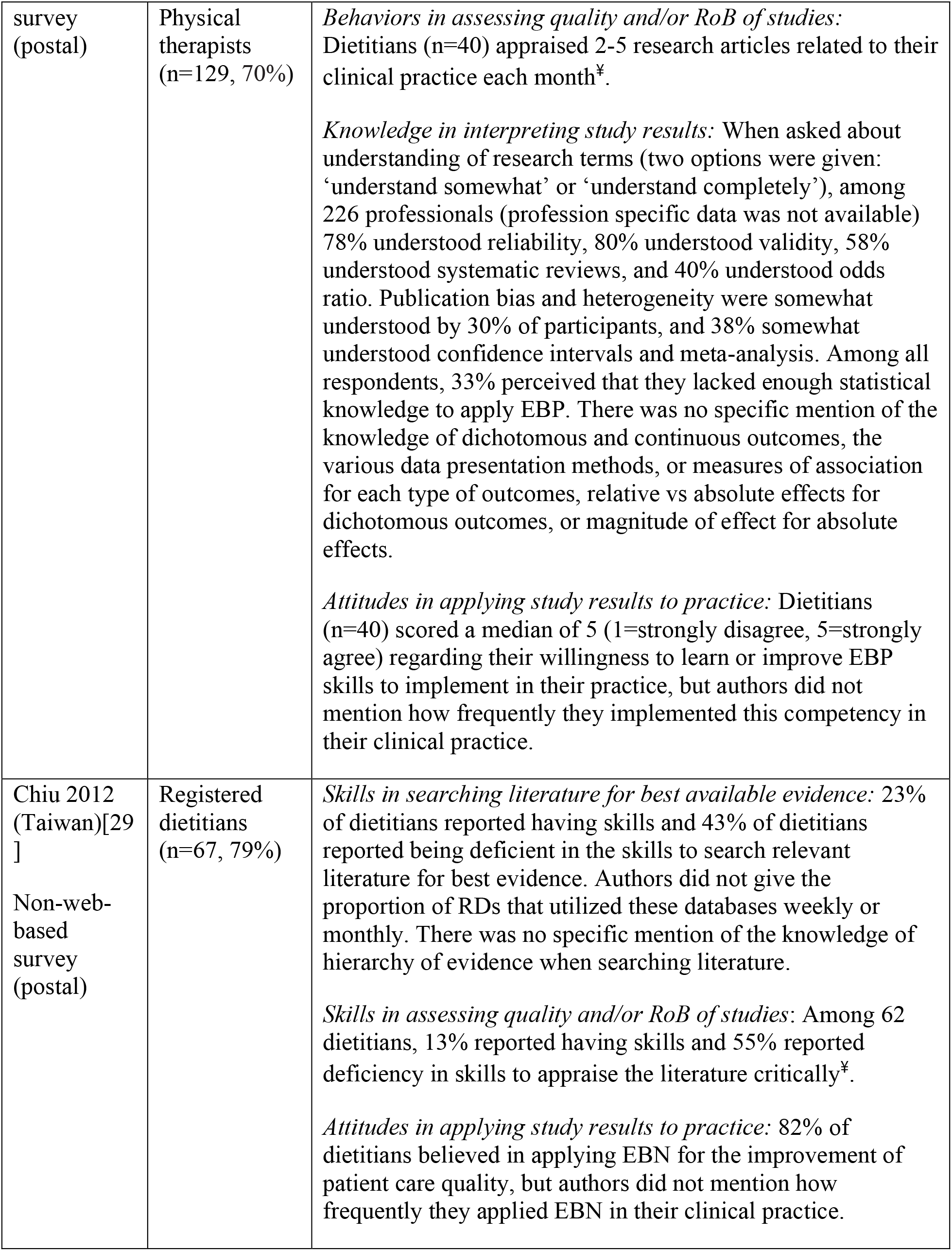
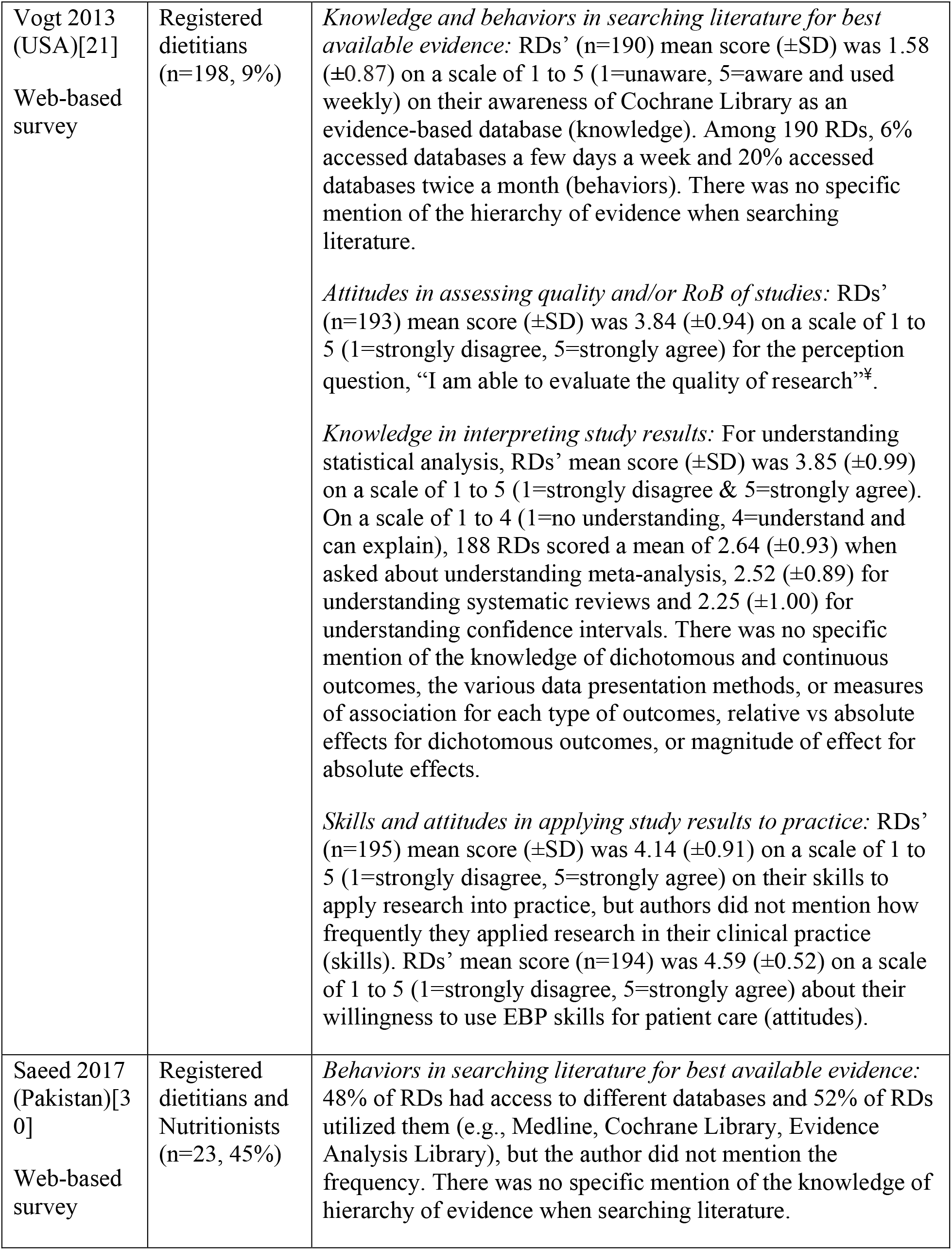
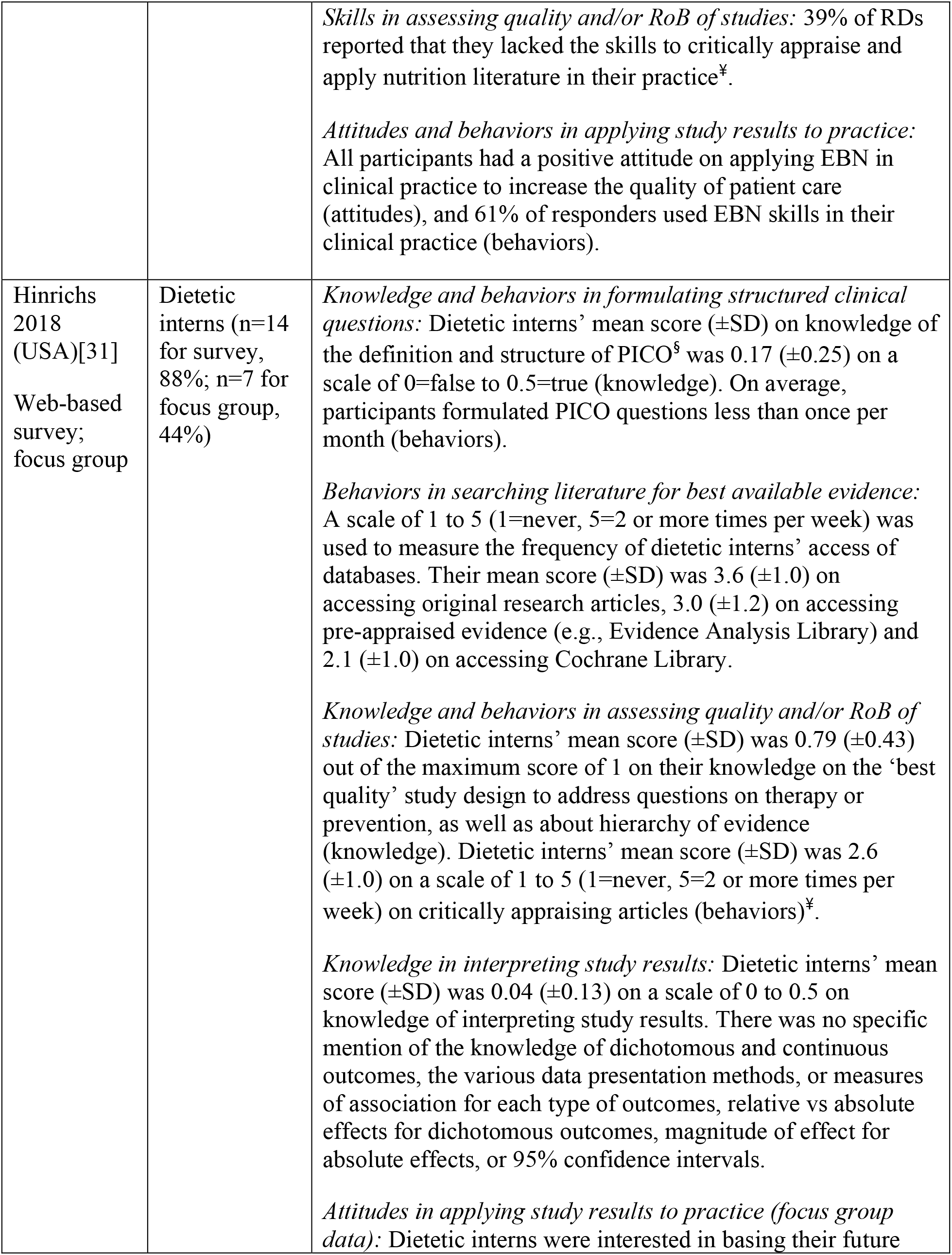
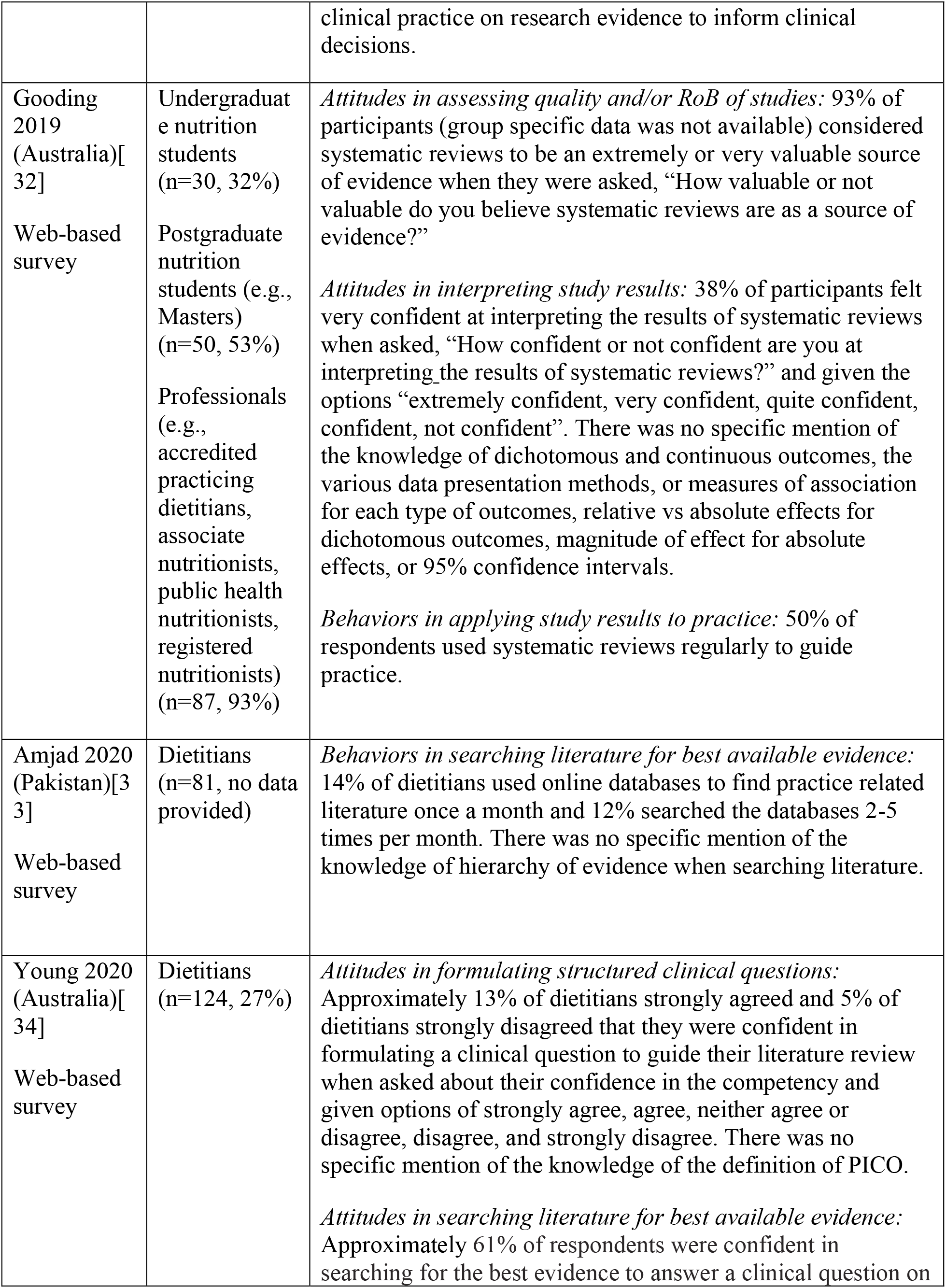
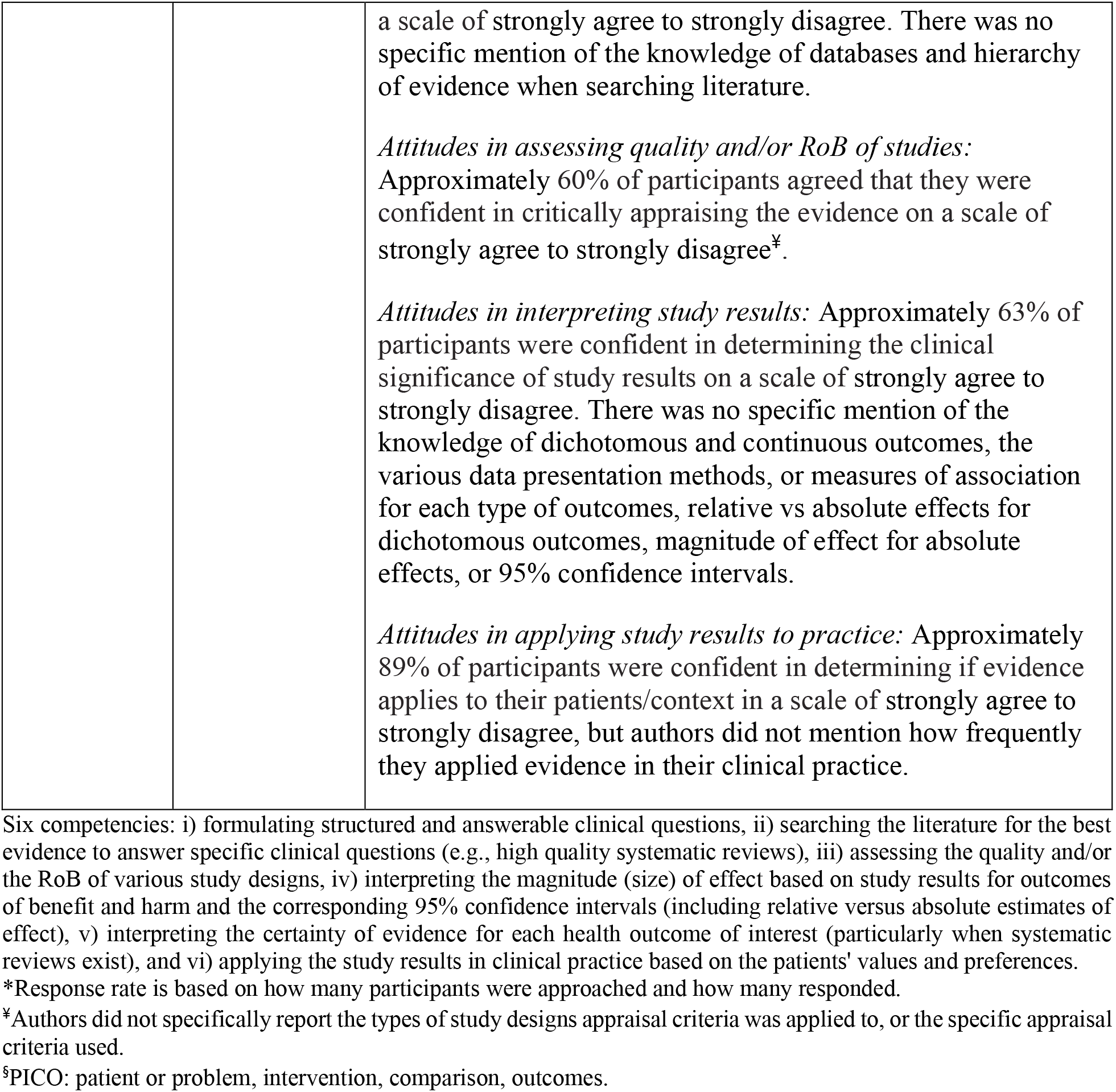
Characteristics of studies documenting EBP competencies.

### Criteria for study inclusion

We included primary studies that assessed knowledge, skills, attitudes, and behaviors related to six EBP competencies among participants (**Table 1**). Eligible participants included clinicians (i.e., registered dietitians (RDs), nutritionists) and nutrition students (i.e., undergraduates, graduates, postgraduates, dietetic interns). Included studies could utilize subjective and/or objective approaches, and report on quantitative or qualitative outcomes. Our target EBP competencies could be measured using questionnaires that were developed by the investigators, adapted from existing instruments, or adopted from already developed instruments such as Fresno test[35] or Evidence-Based Practice Questionnaire[36]).

### Study selection

Our search results were uploaded into a Microsoft Excel spreadsheet (2007) and any study duplicates were removed. Following the guidance from the Cochrane Handbook, two authors, independently and in pairs, screened the titles and abstracts and the full text articles.

### Data extraction

#### Study and participant characteristics

We extracted data, independently and in pairs, from all eligible articles including authors’ last name, publication year, country or region of publication, study design, population characteristics (e.g., profession, education level), EBP outcomes (i.e., knowledge, skills, attitudes, behaviors), EBP competencies, and the detailed characteristics of EBP competency questions from available survey questionnaires (e.g., formulating answerable questions, assessing RoB) including their response options (e.g., Likert scale, multiple choice, dichotomous questions, qualitative data input).

#### Risk of bias assessment

Two reviewers independently assessed the RoB of each cross-sectional observational study (i.e., quantitative survey studies, both web-based and non-web-based, and qualitative study (i.e., focus group)). Risk of bias factors assessed were but not limited to response rate, missing data, clinical sensibility of survey, data collection, data analysis, and clarity of findings. For survey studies, we used a modified version of the CLARITY instrument that included an additional question on the use of sensitivity or subgroup analysis for potential confounding factors[37]. For each question, the instrument uses four response options: ‘definitely low RoB’, ‘probably low RoB’, ‘probably high RoB’ and ‘definitely high RoB’[38]. For focus group studies, we used the Critical Appraisals Skills Programme (CASP) instrument to assess the RoB[39] with three response options: ‘low RoB’, ‘intermediate RoB’ and ‘high RoB’.

#### Quality of reporting assessment

With respect to the quality of study methods reporting, we utilized the CROSS[40] (Consensus-based Checklist for Reporting of Survey Studies) instrument for quantitative studies, and the COREQ[41] (Consolidated criteria for Reporting Qualitative studies) instrument for qualitative focus group studies to assess how comprehensively authors reported population characteristics, study design, data analysis methods and study findings. Two reviewers independently categorized the reporting for each item as: a) clearly reported, b) partially reported, c) unclearly reported, and d) not reported.

#### Questionnaire characteristics and type of competency outcome measured

We extracted the characteristics of questionnaires that were used to assess EBP competencies. We categorized the questionnaires as: a) self-developed (if investigators developed survey questionnaires de novo), b) adapted (if investigators altered existing questionnaires before using them to suit their own study objectives), and c) adopted (if investigators used existing questionnaires verbatim). We also looked at how each study presented the questions from the instruments and categorized them as: a) clearly reported questions, b) partially reported questions, and c) unclearly reported questions. Further, we categorized the competencies assessed by the questionnaires as: a) self-perceived (when participants reported their self-assessment of EBP competencies)[42], and b) objectively assessed (when instruments objectively measured participants’ EBP competencies)[43]. If it was not clear from the study reports, we contacted the authors and asked them to provide their full questionnaire.

### Data analysis

We report our findings descriptively under study and population characteristics, RoB of studies, quality of reporting, and characteristics of the EBP competency questionnaires, while documenting if competencies were self-perceived or objectively (e.g., written or multiple-choice answers) assessed. We could not conduct meta-analysis due to heterogeneous participant groups and methods used to assess the EBP competencies. There was considerable heterogeneity in the questions asked by each study to measure the competencies (e.g., dichotomous, multiple-choice, open-ended questions), as well as variability in reporting central tendency and variance (e.g., some studies used dichotomous response options to calculate proportions, some used means with standard deviations or interquartile ranges).

## RESULTS

### Study and participant characteristics

Our search yielded 2265 initial references. After deduplication, 2002 titles and abstracts were available for screening and 1959 were excluded, leaving 43 full text articles for full text screening. We ultimately included 12 studies that were published between 2001 and 2020[21,24–34], with studies having enrolled between 14 to 258 participants. Details are shown in **Table 1**. All studies reported quantitative survey data (i.e., 7 web-based surveys; 5 non-web-based surveys) with one of the studies[31] also reporting qualitative data (i.e., focus group). Our screening results are outlined in **Figure 1**.

**Figure 1.**
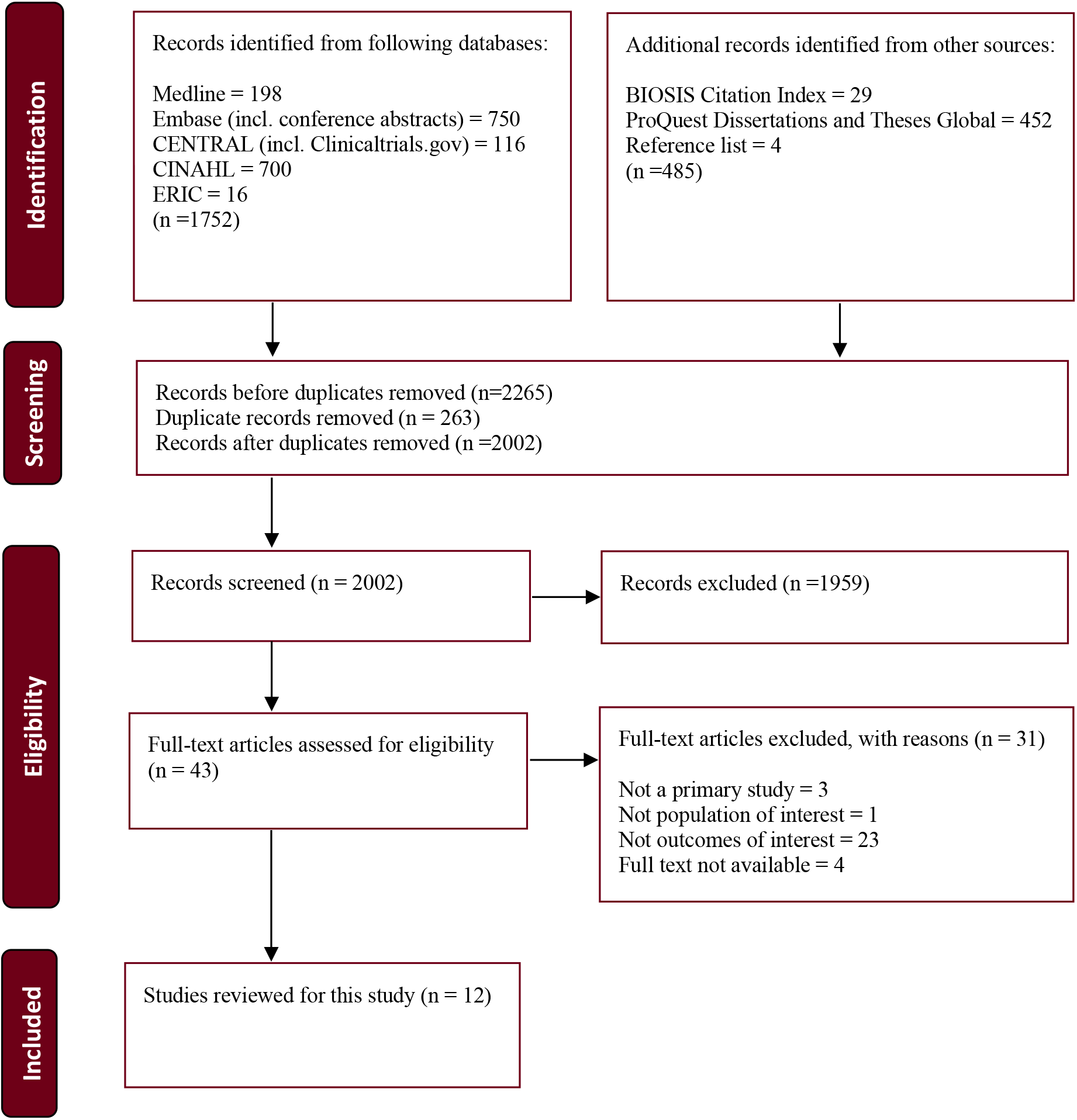
Outline of search strategy depicted by PRISMA Flow Diagram.

Eligible studies were comprised of 1065 participants across six countries (USA, UK, Sweden, Pakistan, Taiwan, Australia). Eleven studies[21,24–30,32–34] included nutrition professionals and one study included dietetic interns[31]. Two studies[30,32] reported enrolling ‘nutritionists’; however, authors did not clarify if nutritionists differed in terms of registration or comprehensive training as compared to RDs. One study[32] reported including undergraduate nutrition students and postgraduate nutrition Masters’ students. Seven studies reported on participants’ age (ranging between 20 years to ≥66 years)[21,25,26,28–30,33], seven studies reported on participants’ education level (i.e., 37% had undergraduate degree and 46% had postgraduate degree)[21,24,26,28–30,32], and 11 studies reported on participants’ employment settings (i.e., 95% were involved in clinical practice and 5% were involved in research)[21,24–30,32–34].

### Questionnaire characteristics

Among 12 cross-sectional survey studies, only three studies (25%) clearly reported the questions they used to measure EBP competencies[28,29,32]. After contacting the authors of nine studies for their full questionnaires, only Hinrichs (2018)[31] was determined to use a questionnaire that, in part, had objective questions on four competencies: formulating structured clinical questions, searching the literature, assessing the quality of studies, and, albeit vaguely, interpreting study results. For information regarding the measurement of EBP competencies on the questionnaires, see **Table 1**.

In terms of the evidence of psychometric properties, five questionnaires had evidence of both reliability and validity, of which two had psychometric testing in RDs[21,26], one had testing in physical therapists[33], and two had testing in an unspecified population[29,31]. Four questionnaires had evidence of validity only, with two reporting validity in an unspecified nutrition population[28,32], and two in an unspecified population[30,34]. One questionnaire had evidence of reliability only, tested in an unspecified nutrition population[24]. Despite some evidence of reliability and validity for our first three competencies, no instruments had explicit questions on determining the magnitude (size) of effect, and none asked about the certainty of evidence for estimates. Further, with respect to applicability, none explicitly asked about the application of patient health-related values and preferences relative to the size and certainty of effect estimates. **Appendix table 1** summarizes the characteristics of the questionnaires.

### Risk of bias and quality of reporting assessment

The overall RoB of included studies varied substantially. Among 12 cross-sectional survey studies, one study was judged as having overall low RoB[26], eight studies had moderate RoB[21,24,25,27–29,31,32], and three studies had high RoB[30,33,34] (**Appendix table2**). The most common RoB issues included no reporting of sensitivity, subgroup, or adjustment analysis for potential confounding factors in eight (67%) studies. The focus group component of Hinrichs (2018)^39^ study was rated as having moderate RoB[31] (**Appendix table3**). Reporting quality from the 40-item CROSS[40] and 32-item COREQ[41] instruments are shown, respectively, in **Figure 2** and **Figure 3**. For details on all items, see **Appendix table4** (CROSS) and **Appendix table5** (COREQ).

**Figure 2.**
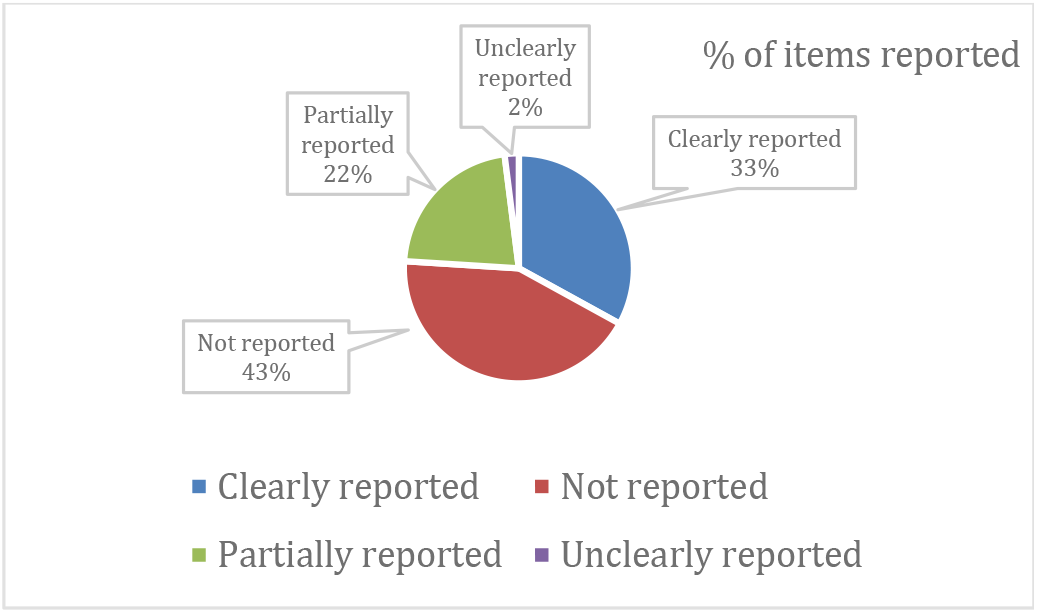
Reporting quality for cross-sectional survey studies.

**Figure 3.**
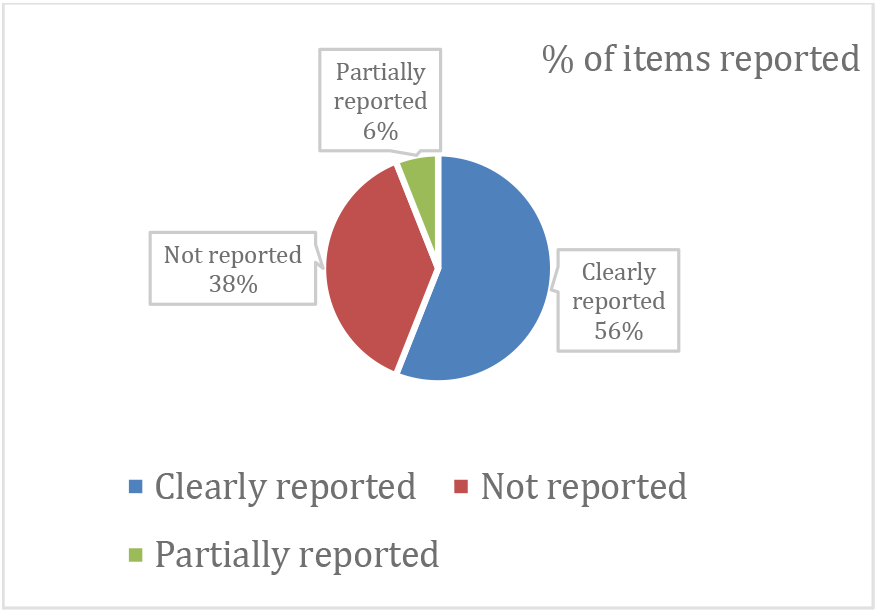
Reporting quality for focus group study.

### Evidence-based practice competencies

#### Formulating structured and answerable clinical questions

Knowledge, attitudes, and behaviors specific to formulating questions was examined by two (17%) studies[31,34]. Hinrichs (2018)[31] asked if dietetic interns (n=14) knew what PICO (patient, population or problem, intervention, comparison, outcomes) referred to, and reported that participants lacked knowledge about the definition and structure of PICO when formulating a clinical question. Young et al (2020)[34] asked dietitians (n=124) to rate their confidence in formulating a clinical question, reporting that only 13% of the participants perceived themselves as confident. Authors did not report how often they performed this competency, or the average number of patients they typically attended to. Regarding behavior, dietetic interns reported formulating PICO questions, on average, less than once per month[31]. Data on the average number of patients over a particular time frame (e.g., per month) was not reported. See **Table 1**.

#### Searching literature for best evidence to answer clinical questions

Knowledge, skills, attitudes, and behaviors specific to searching literature was reported in 10 (83%) studies[21,24–26,28–30,32–34]. Vogt et al (2013)[21] looked at RDs’ (n=190) awareness (knowledge) of different databases containing evidence (e.g., Cochrane Library, Evidence Analysis Library, Medline) and reported that RDs’ awareness was low for the best source of summary data (i.e., Cochrane Library). Thomas et al (2003)[25] and Chiu et al (2012)[29] assessed participants’ skills in searching the literature[25,29] and reported that 81% of dietitians (n=59) and 43% of dietitians (n=67), respectively, lacked skills in searching databases (e.g., Medline) to inform clinical practice. Two studies reported on participants’ attitudes with respect to searching the literature[24,34]. Metcalfe et al (2001)[24] reported that 24% of RDs (n=45) did not feel that searching the literature was a high priority activity, while Young et al (2020)[34] reported that 61% of RDs (n=124) were confident in searching the literature to answer a clinical question. In both study reports there was no specific evaluation of RDs’ knowledge of databases or the hierarchy of evidence when searching the literature. Seven studies reported on the typical behaviors of participants when searching the literature (e.g., percentage and frequency of respondents who searched the literature, which databases were searched)[21,25,26,28,30,31,33]. In six studies (n=629)[21,25,26,28,31,33], 26% of respondents searched the literature at least twice per month (data on average number of patients per month was not reported). The remaining study by Saeed 2017[30] reported that 52% of RDs searched different databases but did not report on the frequency of searching. See **Table 1**.

#### Assessing quality and/or the RoB of evidence based on different study designs

Knowledge, skills, attitudes, and behaviors specific to assessing the methodological quality of the studies was reported in nine (75%) articles[21,24,25,27–31,34], with most reports using the term ‘critical appraisal’ instead of more specific terms such as methodological quality or RoB. For this competency, almost all studies did not report the specific questions (e.g., on selection bias, attrition bias) used to appraise selective study designs. Further, it was often unclear from the study reports if investigators measured participants’ competency in assessing RoB for specific study designs (e.g., systematic review with meta-analysis, randomized controlled trials, cohort), or something else.

Two studies assessed participants’ knowledge on the ‘best quality’ study design[25,31]. Thomas et al (2003)[25] reported that 52% of RDs (n=59) considered clinical practice guidelines, 19% considered systematic reviews, 17% considered local experts, textbooks or case reports, and 12% of RDs considered randomized trials to be the best source of information to address questions on therapy or prevention. Hinrichs (2018)[31] reported that dietetic interns (n=14) had moderate knowledge of the study designs and the hierarchy of evidence. Studies by Metcalfe et al (2001)[24], Thomas et al (2003)[25] and Chiu et al (2012)[29] assessed RDs skills in critical appraisal of scientific literature and reported that, respectively, 60% of RDs (n=27), 86% of RDs (n=51) and 55% of RDs (n=62) lacked the skills. Study by Upton and Upton (2006)[27] reported that RDs’ (n=20) had moderate skills in critical appraisal skills; however, authors and available questionnaires used in the study did not define what was meant by critical appraisal.

Four studies reported on participants’ attitudes regarding, again, undefined critical appraisal of the literature[21,25,32,34]. Among these studies, Thomas et al (2003)[25] found that when participants (n=59) evaluated articles, 93% of them considered study designs as an important criterion for study quality, and 51% of them considered critical appraisal criteria published in the Users’ Guides to the Medical Literature[10] or EBM textbooks. Gooding et al (2020)[32] found that 93% of participants (n=167) perceived systematic reviews to be an extremely or very valuable source of evidence, and Young et al (2020)[34] reported that 60% of RDs (n=124) perceived themselves as confident in critically appraising the evidence.

Two studies reported on participants’ behaviors related to undefined critical appraisal of the literature (e.g., percentage and frequency of participants who appraised the literature to inform clinical practice). Heiwe et al (2011)[28] reported that RDs (n=40) appraised 2 to 5 research articles per month, while Hinrichs (2018)[31] reported that dietetic interns (n=14) performed a moderate amount of appraisal activities to help inform practice. See **Table 1**.

#### Interpreting magnitude (size) of effect and corresponding precision of 95% confidence intervals

Knowledge and attitudes specific to interpreting the study results was discussed in seven (58%) studies[21,24,26,28,31,32,34]. Five studies reported on participants’ overall knowledge in this competency[21,24,26,28,31]. For instance, Metcalfe et al (2001)[24] assessed RDs’ (n=45) and Hinrichs (2018)[31] assessed dietetic interns’ (n=14) understanding or knowledge in interpreting study results. Metcalfe (2001) reported that among 78% of RDs, the statistical analyses in research papers were not understandable, while Hinrichs (2018) reported that all dietetic interns had inadequate knowledge in this competency. Three studies[21,26,28] evaluated participants self-perceived knowledge of statistical terms rather than their actual knowledge of common relative (e.g., risk, odds or hazard ratios) and absolute (e.g., risk difference, number needed to treat) estimates of effect and corresponding estimates of precision (e.g., 95% confidence interval). For instance, Byham-Gray et al (2005)[26] asked about participants’ (n=258) knowledge of the terms ‘relative risk’ and ‘absolute risk difference’ and found that, respectively, 31% and 30% of RDs perceived that they understood or had knowledge of the terms.

Young et al (2020)[34] and Gooding et al (2020)[32] reported on participants’ attitudes towards this competency. Young (2020) examined participants’ (n=124) confidence in determining the clinical significance of study results and 63% of participants reported confidence in their skills. Gooding (2020) asked participants (n=167) “How confident or not confident are you at interpreting the results of systematic reviews?” and reported that 38% felt confident to very confident. No study explicitly reported on participants’ skills to interpret the magnitude (size) of the estimate of effect (e.g., from a trivial to a small, moderate, and large effect) and the corresponding precision of the 95% confidence intervals, nor did they report assessing participants skills in interpreting relative and absolute estimates of effect. See **Table 1**.

#### Interpreting certainty of evidence for each health outcome of interest

While Gooding et al (2020)[32] queried participants about interpreting the results from systematic reviews, no study examined competencies in interpreting the certainty of evidence (e.g., evaluation of consistency of evidence, assessment of publication bias) to support estimates of effect for outcomes of benefit or harm (particularly in the context of systematic reviews with meta-analysis).

#### Applying study results in clinical practice based on patients’ values and preferences

Skills, attitudes, and behaviors in applying study results in clinical practice were heterogeneously assessed in nine (75%) studies[26–32,34,44]. Vogt et al (2013)[21] and Upton and Upton (2006)[27] examined participants’ skills in applying study results to their practice and reported that participants perceived themselves as skilled in this competency. Eight studies reported on participants’ attitudes or willingness in applying study results in practice, of which studies by Vogt et al (2013)[21], Heiwe et al (2011)[28] and Upton and Upton (2006)[27] reported that participants’ (n=254) had a moderate to high degree of willingness towards this competency. Studies by Chiu et al (2012)[29], Hinrichs (2018)[31] and Saeed (2017)[30] reported that participants either believed in (82% among 67 participants), were interested in (100% of 7 participants), or had a positive attitude toward (100% of 23 participants) applying study results. The remaining two studies by Young et al (2020)[34] and Byham-Gray et al (2005)[26] reported that participants perceived themselves, respectively, as confident (89% among 124 participants) and capable of (most or all of 258 participants, no proportion was given) applying evidence in their practice. Two studies[30,32] reported on participants’ behaviors in applying study results among which Gooding (2020)[32] reported that 50% of the respondents (n=167) used systematic reviews regularly to guide their practice.

Applicability was vague across all studies. For instance, it was unclear if investigators looked at participants’ competency in applying the best available evidence (e.g., high quality systematic reviews with meta-analysis) based on estimates of benefits and harms of an intervention with patients, or something else. Moreover, no studies explicitly reported on applying patients’ values and preferences based on the best available evidence. See **Table 1**.

## DISCUSSION

### Summary of findings

Our systematic review of EBP competencies included 12 cross-sectional surveys comprised of 1065 participants (i.e., RDs, nutritionists, dietetic interns, and nutrition students) across six countries. The overall reporting quality among the surveys was poor, with only 33% of items clearly reported and the survey questions were predominantly self-perceived assessments. There were also considerable deficiencies across studies regarding the measurement of EBP competencies. For instance, the six competencies were often incompletely defined or reported (e.g., it was unclear what ‘applicability’ and ‘critical appraisal’ referred to, and what study designs were appraised by the participants), which made it difficult to compare studies and to reach an overall conclusion. Further, no studies had explicit questions on two (33%) of the six core EBP competencies (i.e., determining the magnitude (size) of effect, determining the certainty of evidence for estimates), and study reports were unclear with respect to competencies in applying patient values and preferences relative to the size and certainty of effect estimates, skills that are essential for optimizing clinical and public health nutrition decision-making.

### Strengths of this study

To our knowledge, this is the first systematic review that has evaluated EBP competencies in the field of nutrition. The competencies assessed are based on the Users’ Guides to the Medical Literature[10] and a consensus statement on EBP competencies for health professionals[23]. In addition to medicine[3], these competencies have generally been embraced in many contemporary evidence-based programs including nursing[5], pharmacy[8], physiotherapy[6], occupational therapy[9], and psychology[7]. We worked with an experienced librarian to conduct a comprehensive search across five databases and four grey literature sources with no language restrictions, and we registered our study protocol on an open-access, publicly accessible website[20]. We performed screening, data extraction, and quality assessment independently and in pairs, including the assessment of our 12 studies using CROSS[40] and COREQ[41] reporting instruments. Finally, we used the PRISMA[18] and SWiM[19] reporting standards to provide a transparent and clear presentation of our findings.

### Limitations of this study

Our study protocol, published *a priori*[20], underwent several revisions that were necessary once we better understood the available data. The first revision was the exclusion of the seventh competency, which involved self-evaluation of EBP competencies. This decision was made because it seemed impractical to expect participants to perform self-evaluation when there appeared to be a limited understanding of the first six competencies. This aspect of limited understanding may have resulted from poor reporting in 12 eligible studies, as determined by CROSS[40] and COREQ[41] assessments, and no reporting on competencies five and six (i.e., effect size, and certainty of evidence). Although there has been a longer list of EBP competencies proposed for medical and allied health practitioners[23], we emphasized six core EBP competencies that are directly related to treatment and prevention[10,23], competencies that are long standing and directly relevant to nutrition and the broader health professions practice. With respect to the assessment instruments, the second departure from our protocol was the inclusion of an analysis of the reporting quality of surveys and focus groups. Post-hoc, we decided to add these assessments given that we were surprised by how poorly the studies documented key items that were used to assess competencies (e.g., clear reporting of the questions regarding assessing the methodological quality or RoB). Finally, we used the CLARITY[45] instrument to assess the RoB across survey studies, an instrument that does not have a peer-reviewed publication, or established evidence of validity and reliability. In keeping with rationale for the development of the CLARITY cross-sectional RoB instrument, we used the tool due to our inability to find a comprehensive instrument that addresses RoB in surveys of attitudes and practices[46]. It should be noted that the response options on the CLARITY instrument[46] are based on the Cochrane RoB instrument which has established evidence of validity and reliability[47].

### Implications for clinical practice

The accreditation standards for dietetic programs give flexibility[13–16] for diverse programs to define where and how to incorporate training in EBP competencies into their curricula which leaves the possibility for heterogeneity in the training, understanding and application of EBP competencies among dietetics practitioners[48]. Since EBP questions are included in the registration examination for dietitians[49–51], our review shows the urgency to offer specific information on EBP competencies through dietetic curricula that nutrition trainees should learn and use in practice (such as locating literature through formulating clinical questions using the PI(E)CO (population/problem, intervention/exposure, comparison, outcome) format; evaluating the available literature for methodological quality or RoB using specific tools for different study designs; interpreting the effect size and precision of effects from the literature). That, in turn, will contribute to the increased utilization of EBP competencies in clinical and public health nutrition settings and serve to benefit patients and dietitians by promoting more fully informed decisions[52].

Although our primary objective was not to systematically collect data on training, we documented it whenever available. Only seven (58%) studies from our review reported on participants’ training in EBP[21,25,26,28,29,32,34]. For instance, Byham-Gray et al (2005)[26] reported that 55% of RDs (n=258) received “critical appraisal training”, and Chiu et al (2012)[29] reported that 27% of RDs (n=67) took an educational course in “evidence-based nutrition”[53]. Considering that EBP is an important skill endorsed by dietetic associations worldwide, it would be beneficial to increase the standardization of EBP training to better support dietitians and to further promote interdisciplinary care. Various pedagogical approaches are available to teach foundational EBP competencies at different levels of education including journal clubs and critical appraisal courses, and educators may select those with proven effectiveness[54]. To build on foundational EBP competencies, integrating case-based learning, which showcases the real-life application of EBP competencies into the current framework of the Nutrition Care Process (i.e., at the time of developing Problem, Etiology and Signs/Symptoms (PES) statements and identifying an appropriate nutrition intervention) would enable dietetic learners to better recognize and appreciate the impact of EBP competencies in clinical decision-making across different domains of dietetics practice[55]. This integration would ultimately assist dietitians in their interactions with patients. Additionally, standardized RD board exam questions that address each of our EBP competencies would promote improved training in various nutrition programs. While it is expected that some patients will be uncomfortable with a detailed, fully enumerated discussion of the absolute estimates and certainty of estimates for the benefits and potential harms for a given nutritional intervention[56], and prefer to leave the decision to the clinician, based on published evidence it is anticipated that many patients will be very comfortable, and relieved to learn of such details, allowing for fully informed decision-making[2,57].

Several studies have shown that there is a growing recognition and reliance on evidence-based clinical practice guidelines in the field of dietetics[21,25,26,28,33]. These guidelines are consistently emphasized in educational curricula and professional standards documents. This highlights the need for awareness and increasing access to practice guidelines that are relevant to different areas of clinical dietetic practice. Nonetheless, we need to keep in mind that not all guidelines are created equal (e.g., many lack adherence to Institute of Medicine 2011 and National Guideline Clearinghouse Extent of Adherence to Trustworthy Standards (NEATS) 2019 criteria). Further, while resources like the Evidence Analysis Library from the Academy of Nutrition and Dietetics provide some evidence-based guidelines, they are not regularly updated, and many of the existing reviews are outdated by more than five years[58]. Therefore, it is essential to develop evidence-based guidelines in accordance with established standards, consistently update them, and offer systematic training to dietitians that would enable them to locate, evaluate, and apply these guidelines. Currently, many guidelines are developed by professional societies outside of dietetics, focusing on specific areas (e.g., cardiometabolic disease risk[59], pediatric nutrition[60]). This requires dietitians to be familiar with these societies, often requiring membership and additional fees to access guidelines. These barriers should be addressed to ensure dietitians’ familiarity with and adherence to the evidence-based guidelines. This will enable clinical practitioners to effectively engage with the literature and apply it to their practice.

### Implications for research

Future cross-sectional surveys should comprehensively assess knowledge, skills, attitudes, and behaviors for core EBP competencies using valid and reliable instruments to more objectively measure each of the competencies[61], in addition to following CROSS[40] reporting instrument. For instance, a well-rounded set of EBP skills should encompass instruction and evaluation in the following areas: 1) understanding research study findings, which involves assessing the magnitude (size) of effects (absolute estimates such as mean difference, risk difference and numbers needed to treat for benefit/harm, and relative estimates such as relative risk, odds ratios, and hazard ratios, along with measures of precision e.g., 95% confidence interval)[62], 2) understanding the certainty of evidence for estimates, particularly for systematic reviews with meta-analysis, and 3) demonstrating proficiency in applying results based on patient values and preferences relative to the size and certainty of effect estimates. These competencies, considered essential to decision-making[63], have not been evaluated in studies to date. Research should be conducted to help elucidate how these competencies may impact satisfaction with the dietitian-patient encounter[57]. Furthermore, it is important to distribute surveys to targeted professional groups that are focused on specific areas of practice (e.g., dietetic practice groups within the Academy of Nutrition and Dietetics[64], or international dietetic societies like Genetic Metabolic Dietitians International[65]). This ensures that accurate data is gathered to gain insights into the implementation of EBP in specific clinical settings. Finally, to improve EBP competencies, research on various teaching strategies (e.g., seminars, workshops, courses, journal clubs) is also required[66,67].

## CONCLUSION

Among 12 included articles, there were considerable deficiencies across studies regarding the measurement of EBP competencies. In addition to the questions being predominantly self-perceived, as opposed to objective assessments, the six competencies were often incompletely defined or reported, which made it difficult to compare studies. No studies reported explicit questions on two (33%) of the six core EBP competencies (i.e., determining the magnitude (size) of effect, determining the certainty of evidence for estimates), skills that are essential for optimizing clinical and public health nutrition decision-making.

## Data Availability

All data produced in the present study are available upon reasonable request to the authors.

## Acknowledgement

We thank Delaney Sauers (undergraduate nutrition student) for helping with our preliminary literature search and article screening, Joshua Goldenberg for assistance with piloting our data extraction forms, and Erin Boyce (Librarian) for her assistance with our initial search for articles addressing EBP competencies.

## Contributors

BCJ conceived the study. NRG and BCJ designed the study. NRG and BCJ developed *a priori* list of core evidence-based competencies for nutrition professionals and students. NRG, BCJ and MC designed and performed the search strategy. NRG and RAC selected the articles. NRG, ZE, JZ, and SGL extracted the data. NRG analyzed the data and wrote the first draft of the manuscript. BCJ reviewed the data of the draft manuscript. All authors critically revised the manuscript for important intellectual content and contributed to the writing of the final version of the manuscript. All authors agreed with the results and conclusions of this article. The corresponding author attests that all listed authors meet authorship criteria and that no others meeting the criteria have been omitted. NRG and BCJ are the guarantors.

## Funding

This research was not funded by a specific grant from any funding agency in the public, commercial, or not-for-profit sectors. It was funded, in small part, by the Presidential Transformational Teaching Grant from Texas A&M University (awarded to BCJ). The university had no role in study design, data collection, data analysis, data interpretation, or writing of the report. The corresponding author had full access to all the data in the study and had final responsibility for the decision to submit for publication.

## Conflict of interest

All authors have completed the ICMJE uniform disclosure form at www.icmje.org/disclosure-of-interest/ and declare: support from Texas A&M University for the submitted work; BCJ received a grant from Texas A&M AgriLife Research to fund investigator initiated research related to saturated and polyunsaturated fats. The grant was from Texas A&M AgriLife institutional funds from interest and investment earnings, not a sponsoring organization, industry, or company.

## Ethical approval

Not required. All the work was developed using aggregate level data.

## Data sharing

Further data is available on request through the corresponding author at.

## Notes

### Competing Interest Statement

The authors have declared no competing interest.

### Clinical Protocols

https://www.crd.york.ac.uk/prospero/display_record.php?ID=CRD42022311916

