## Supplementary materials for "Evidence-Based Practice Competencies among Nutrition Professionals and Students: A Systematic Review"

Nirjhar R. Ghosh, MSc, doctoral student; Zahra Esmaeilinezhad, MSc, doctoral student; Joanna Zając, PhD, Assistant Professor; Rebecca A. Creasy, PhD, NSCA-CPT, Instructional Assistant Professor; Saundra G. Lorenz, MS, RD, LD, Senior Lecturer; Molly Crews, MLS, Systematic Review Librarian; Karen M. Beathard, PhD, RD, LD, FAND, Instructional Associate Professor; Malgorzata M. Bala, MD, PhD, Profesor; Kevin C. Klatt, PhD, RD, Assistant Research Scientist; Bradley C. Johnston, PhD, Associate Professor

Appendix A. Search strategy: Medline
Appendix Table 1. Characteristics of instruments used to measure EBP competencies
Appendix Table 2. Assessment of risk of bias for cross-sectional survey studies
Appendix Table 3. Assessment of risk of bias for focus group
Appendix Table 4. Percentage of CROSS reporting criteria satisfied by individual studies
Appendix Table 5. Percentage of COREQ reporting criteria satisfied by studies

### Appendix A

### Search strategy: Medline (1946 - March 10, 2023)

| | |
|---|---|
| 1 | (nutrition adj2 (student* or trainee* or professional* or intern* or specialist or undergrad* or grad*)).tw,kw,ti,kf. (1846) |
| 2 | limit 1 to last 2 years (329) |
| 3 | (dietetic intern* or registered dietitian* or registered dietician* or registered nutritionist or nutritionist* or dietitian* or dietician*).tw,kw,ti. (12781) |
| 4 | limit 3 to yr="2022 -Current" (1319) |
| 5 | ((certified or clinical) adj2 nutrition specialist*).tw,kw,ti,kf. (9) |
| 6 | limit 5 to yr="2022 -Current" (1) |
| 7 | exp Nutritionists/ or exp Dietetics/ (9207) |
| 8 | limit 7 to yr="2022 -Current" (250) |
| 9 | 1 or 3 (14386) |
| 10 | 1 or 3 or 5 or 7 (20309) |
| 11 | (evidence-based practice or EBP or evidence based practice or evidence-based or evidence based or evidence-based nutrition or evidence based nutrition or EBN or evidence-inform* or evidence inform*).tw,kw,ti. (158196) |
| 12 | limit 11 to yr="2022 -Current" (18506) |
| 13 | ((EBP or evidence-based or evidence based) adj3 (principle* or framework* or competenc* or decision or healthcare)).tw,kw,ti,kf. (6147) |
| 14 | limit 13 to yr="2022 -Current" (766) |
| 15 | exp Evidence-Based Practice/ or exp Evidence-Based Medicine/ or exp Translational Medical Research/ (105353) |
| 16 | limit 15 to yr="2022 -Current" (1285) |
| 17 | knowledge translation.tw,kw,ti. (4228) |
| 18 | limit 17 to yr="2022 -Current" (546) |
| 19 | 11 or 13 or 15 or 17 (227333) |
| 20 | (knowledge or skill* or attitude* or behavior*).tw,kw,ti. (2278182) |
| 21 | limit 20 to yr="2022 -Current" (220843) |
| 22 | exp Knowledge/ or exp Health Knowledge, Attitudes, Practice/ or exp Clinical Competence/ or exp Professional Competence/ or exp Attitude/ or exp Behavior/ (2403032) |
| 23 | limit 22 to yr="2022 -Current" (91141) |
| 24 | 20 or 22 (4044302) |
| 25 | (survey* or questionnaire* or interview* or cross-sectional or cross sectional or focus group*).tw,kw,ti. (1920847) |
| 26 | limit 25 to yr="2022 -Current" (216874) |
| 27 | exp Surveys/ or Questionnaires/ or exp Cross-Sectional Studies/ (1506932) |
| 28 | limit 27 to yr="2022 -Current" (85828) |
| 29 | 25 or 27 (2613814) |
| 30 | 10 and 19 and 24 and 29 (198) |

### Appendix Table 1. Characteristics of instruments used to measure EBP competencies

| Reference (country)<br><br>Study design | Characteristics of questionnaires used to measure EBP competencies | Question reporting category (clearly reported, partially reported, unclearly reported) |
|---|---|---|
| Metcalfe 2001 (UK)[24]<br><br>Survey | Authors adapted *Barriers and Attitudes to Research in the Therapies* questionnaire from previous studies[44,45]. The original questionnaire was developed for dietitians, occupational therapists, physiotherapists and speech and language therapists. Author questionnaire had 3 sections:<br>1. Demographic details.<br>2. Perceived importance of research (PIR) section, consisting of 7 questions (agreement with a statement scored as -1 and disagreement as +1).<br>3. Perceived barriers (PB) section, consisting of 22 questions (agreement with a statement scored as –1 and disagreement as +1). | Partially reported. Authors mentioned the questions from section 1 in table 1, section 2 (PIR) in table 2, and section 3 (PB) in table 3. All the questions were without their full form, and the authors did not provide the questionnaire as a supplement.<br><br>It was not clear what authors meant by statistical analyses (if they asked about understanding p-value, 95% CI, RR, ARR), and how they measured evaluation of research (did they mean assessing RoB or quality of study or certainty of evidence). |
| Thomas 2003 (Australia)[25]<br><br>Survey | Authors adapted the questionnaire from Scott et al (2000)[46] that was originally developed for physicians, and it had 6 sections:<br>1. Demographic details.<br>2. Identifying information needs (options included <5, ≥5 and ≥10 times week).<br>3. Searching the literature and electronic databases (options included <5, 5–10, or >10).<br>4. Critical appraisal (the question was stated as *Criteria used by dietitians to assess study quality*, followed by several options such as *the journal in which the article was published; the authors of the study; the institution where the study was performed; the objectives of the study; the study design; the global impression of the validity of the data; the validity/quality based on objective critical appraisal criteria (e.g. those published in the Users' guides to the literature or EBM textbooks); the opinions of colleagues in relation to the article; the similarity of the results to the recommendations of existing hospital guidelines or any other criteria (to be specified)*).<br>5. Perceived barriers to applying EBN (Options included *lack of time; difficulties in searching for the articles they needed (e.g. defining the search strategy, accessing electronic databases, accessing journals or culling articles with irrelevant content); lack of skills for evaluating the quality of the articles (e.g. assessing study methods, understanding statistics or applying critical appraisal criteria) and difficulties in applying information to their patients (e.g. their patient population was too dissimilar from that in published studies; there were economic constraints; personal considerations such as their own sense of caution and* | Partially reported. Authors mentioned section 4 questions (critical appraisal) in table 1, but they did not mention the questions for the remaining five sections. Authors did not provide the questionnaire as a supplement, and it was not clear what they meant by understanding statistics (if they asked about understanding p-value, 95% CI, RR, ARR). |

| | | |
|---|---|---|
| | *conservatism; difficulty applying results that were at odds with their own experience; or resistance, e.g. from other health professionals to proposed changes to practice) and other (to be specified)*).<br>6. Acquiring or teaching EBN skills (They were asked to rank on a scale of 1 (strongly believe) to 5 (disbelieve), the extent to which they believed in the philosophy and principles of EBN). | |
| Byham-Gray 2005 (USA)[26]<br><br>Survey | Authors developed a questionnaire entitled *Dietitian Research Involvement* with 3 sections:<br>1. 21 items that measured attitudes and knowledge of research using a modified version of the 29-item *Barriers to Research Utilization Scale*[47]. Response options used a 5-point Likert scale: 1=strongly agree, 5=strongly disagree.<br>2. 17 items addressing specific areas of knowledge, behaviors about knowledge, attitudes towards EBP, incorporating portions of the instrument entitled *General Practitioners' Perceptions of the Route to Evidence-Based Medicine*[48]. Response options: 5-point scale (1=strongly disagree, 5=strongly agree), 4-point scale (1=unaware and 4=use), and 3-point scale (1=don't understand and 3=do understand). Coded dichotomous questions were scored as (1=no, 2=yes).<br>3. Socio-demographic details. | Partially reported. Authors mentioned section 1 questions in table 2, section 2 questions in table 3, and section 3 questions in table 1. It was unclear how to map 21 items from section 1 and 17 items from section 2 to the questions presented in tables 2 and 3, respectively. All the questions were without their full form, and the authors did not provide the questionnaire as a supplement.<br><br>It was not clear what authors meant by training in search strategy (if they meant formulating clinical questions using PICO format, search using Boolean), understanding statistical analysis (if they asked about understanding p-value, statistical tests) and critical appraisal (assessing study quality or RoB). |
| Upton 2006 (UK)[27]<br><br>Survey | Authors adapted the questionnaire from Upton and Lewis[49] which originated for healthcare professionals overall. The current questionnaire had 4 sections:<br>1. Knowledge of clinical effectiveness (CE) and EBP. In this section, a visual analogue scale was used to rate participants' knowledge. A scale of 1 to 7 (the lower the 'poorest' and the higher the 'best') was used to rate skills on EBP.<br>2. Practice of Individual Components of CE and EBP. Here, the frequency of completing the key steps of EBP was rated from 'never' to 'frequently'. Individuals also rated their likelihood of acting on evidence on a 5-point Likert type scale (5=most likely).<br>3. Barriers to the Implementation of EBP section used a five-point scale (with increasing scores indicating increasing difficulty).<br>4. Demographic details. | Partially reported. Authors mentioned section 1 questions in table 2 ('Knowledge of clinical effectiveness' part) and table 3 ('Skill ratings' part). A part of the section 2 questions was mentioned in table 4 ('Likelihood of acting on evidence' part), but no questions were mentioned for 'Practice of Individual Components' part. Section 3 and 4 questions were mentioned in tables 5 and 1, respectively. All the questions were without their full form, and the authors did not provide the questionnaire as a supplement.<br><br>It was not clear if authors meant formulating questions using PICO format, tracking down evidence using Boolean operators. They did not clarify what they meant by critical appraisal or determining validity of study (assessing study quality or RoB, determining certainty of evidence), or what was the |

| | | |
|---|---|---|
| | | standard to evaluate one's own practice. |
| Heiwe 2011 (Sweden)[28]<br><br>Survey | Authors adopted a questionnaire from Jette et al (2003)[50] that was originally created to assess the attitudes, beliefs and knowledge of physical therapists towards EBP. The questionnaire comprised 5 sections and 51 statements:<br>1. 14 items on attitudes towards the use of evidence, perceived benefits, and limitations of evidence-based practice.<br>2. 6 items on the use and understanding of clinical practice guidelines.<br>3. 10 items on the availability of resources to access information and the skills required for the use of those resources.<br>4. 2 items regarding the understanding of research terms and perceived barriers to evidence-based practice.<br>5. 19 items on demographic details.<br><br>Majority of items were scored on a five-point Likert scale (1=strongly disagree to 5=strongly agree). | Clearly reported. Authors mentioned all the questions in table 2 and appendix A. |
| Chiu 2012 (Taiwan)[29]<br><br>Survey | Authors adapted a questionnaire based on existing questionnaires that originated for measuring physicians' usage of online database[51], and physicians and nurses' views on EBP[52]. The current questionnaire included items on:<br>1. Demographic data.<br>2. Patterns of information-searching. The patterns were rated by a 5-point Likert scale (always, often, sometimes, seldom, and never).<br>3. Awareness of, belief in, attitude toward, knowledge of, and skill in evidence-based nutrition (EBN). Questions regarding the belief in, attitude toward, knowledge of, and skill in were rated by a 5-point Likert scale (strongly agree, agree, neutral, disagree, and strongly disagree).<br>4. Barriers to implementing EBN. The questions were rated on a 5-point Likert scale (strongly agree, agree, neutral, disagree, and strongly disagree). | Clearly reported. Authors mentioned section 1 questions in tables 2 and 3; section 2 questions in tables 4, 5 and 6; and sections 3 and 4 questions, respectively, in figures 1 and 2. |
| Vogt 2013 (USA)[21]<br><br>Survey | Authors adapted *Dietitian Research Involvement* instrument from Byham-Gray et al (2005)[26] which was originated for measuring dietitians' research involvement, and included questions on:<br>1. Demographic details.<br>2. Clinical use of EBP. Frequency of clinical use of EBP resources was measured via a 5-point Likert scale (5=a few days a week to 1=never); attitudes questions were rated via a 5-point Likert scale (5=strongly agree to 1=strongly disagree).<br>3. Perceptions, attitudes, and knowledge of EBP. Perceptions and attitudes questions were rated via a 5-point Likert scale (5=strongly agree to 1=strongly | Partially reported. Authors mentioned section 1 questions in table 1, section 2 questions in tables 2 and 3, and section 3 questions in table 4. All the questions were without their full form, and the authors did not provide the questionnaire as a supplement. It was not clear what they meant by understanding statistical analysis (if they asked about understanding p-value, statistical tests) and critical |

| | | |
|---|---|---|
| | disagree). Knowledge questions were rated via a 5-point Likert scale (5=aware and used weekly to 1=unaware), a 4-point scale (4=understand and can explain to 1=no understanding) or dichotomous questions (2=yes to 1=no). | appraisal (assessing study quality or RoB). |
| Saeed 2017 (Pakistan)[30]<br><br>Survey | Authors developed a structured questionnaire to explore the knowledge and use of EBP among dietitians and nutritionists. Where most of the responses were multiple choice questions with a few open-ended questions. No other details were given about the questionnaire. | Unclearly reported. Authors mentioned the questions related to knowledge and the use of EBP in table 2. All the questions were without their full form, and the authors did not provide the questionnaire as a supplement. It was not clear which questions were multiple choice and which questions were open-ended. |
| Hinrichs 2018 (USA)[31]<br><br>Survey, focus group | For the survey part of the study, the author adapted the questionnaire from Vogt et al (2016)[53] which originated to measure RDs' EBP knowledge and clinical practice behavior. The questionnaire had two sections:<br>1. The first section assessed EBP knowledge (measured by free-text (2-point), multiple choice questions (1-point), and true/false questions (0.5-point)).<br>2. The second section assessed the clinical practice behavior, measured by how often EBP behaviors were performed. The scoring system for this section was modified to use a 5-point Likert scale (5=2 or more times per week; 4=once per week; 3=one to 3 times per month; 2=less than once per month; 1=never)).<br><br>For the focus group part of the study, the author developed questions to guide the discussion and asked follow-up questions during the focus group to delve deeper into participants' responses. | For survey: partially reported. The author mentioned section 1 questions in table 1 and section 2 questions in table 2. All the questions were without their full form, and the authors did not provide the questionnaire as a supplement.<br><br>For focus group: clearly reported. The author provided the focus group questionnaire as a supplement. |
| Gooding 2019 (Australia)[32]<br><br>Survey | Authors adapted the questionnaire from previous studies exploring the perceptions of EBP[26] and perceptions of research[54] in nutrition professionals. It included questions on:<br>1. Participant demographics.<br>2. Research experience.<br>3. Perceptions of, use of, barriers to use and conduct of systematic reviews (SRs).<br>4. Knowledge of SRs.<br>5. Training and feedback to increase SR use. | Clearly reported. Authors mentioned all the questions in the supplemental table S1. |
| Amjad 2020 (Pakistan)[33]<br><br>Survey | Authors adopted a questionnaire from Jette et al (2003)[50] which was developed to assess the beliefs, attitudes, knowledge and behaviors of physical therapists towards EBP. The original questionnaire was comprised of 31 items to gather information about:<br>1. Interest and inspiration to involve in EBP.<br>2. Expertise related to retrieving and interpreting information. | Unclearly reported. Authors did not mention the questions from the original questionnaire that were addressed in tables 1, 2 and 3. They used the titles of the tables as "EBP is necessary in practice", "Read related literature" and "Use databases for literature search" but it was unclear how to map the titles to any |

| | | |
|---|---|---|
| | 3. Skill to access information.<br>4. Access and usage of practice guidelines.<br>5. Barriers in using EBP. | of the five sections of the original questionnaire. It was not clear if the authors used all 31 items from the original questionnaire or only three items. They did not provide the questionnaire as a supplement. |
| Young 2020 (Australia)[34]<br><br>Survey | Authors developed a novel questionnaire based on existing questionnaires wherever possible. The novel questionnaire contained four sections:<br>1. Participant characteristics.<br>2. Familiarity with KT including previous training.<br>3. Confidence undertaking KT process (e.g., identifying problems, reviewing or adapting knowledge or evidence, assessing barriers, selecting or implementing interventions, monitoring, evaluating, and sustaining knowledge in practice).<br>4. KT training and support preferences and barriers to participate in training. | Unclearly reported. Authors mentioned section 1 questions in table 1 and section 3 questions in figure 1. They did not mention the questions from section 2 and 4. They also did not cite the original questionnaires used in developing the current questionnaire. They mentioned a ProQuest link as a source of the final questionnaire which was not accessible: https://dcjournal.ca/doi/suppl/10.3148/cjdpr-2019-027/suppl_file/cjdpr-2019-027suppla.docx#supplementary-materials |

### Appendix Table 2. Assessment of risk of bias for cross-sectional survey studies

| Reference | Q1 | Q2 | Q3 | Q4 | Q5 | Q6 |
|---|---|---|---|---|---|---|
| Metcalfe 2001[24] | | | | | | |
| Thomas 2003[25] | | | | | | |
| Byham-gray 2005[26] | | | | | | |
| Upton 2006[27] | | | | | | |
| Heiwe 2011[28] | | | | | | |
| Chiu 2012[29] | | | | | | |
| Vogt 2013[21] | | | | | | |
| Saeed 2017[30] | | | | | | |
| Hinrichs 2018[31] | | | | | | |
| Gooding 2019[32] | | | | | | |
| Amjad 2020[33] | | | | | | |
| Young 2020[34] | | | | | | |

**Risk of bias criteria for survey studies:** If any two of the items (i.e., sources of population, response rate, missing data within questionnaire, clinical sensibility of survey, validity of survey instrument, sensitivity analysis for potential confounding) had high RoB, the study was deemed high RoB. If a study had only one high risk item, it was deemed moderate RoB, and if it had no high-risk item, it was considered low RoB. **Questions:** Q1. Is the source population representative of the population of interest? Q2. Is the response rate adequate? Q3. Is there a little missing data? Q4. Is the survey clinically sensible? Q5. Is there any evidence for the validity of the survey instrument? Q6. Was there an assessment of, or sensitivity analysis for potential confounding for the main study results? Green indicates an item as having low RoB, yellow indicates moderate RoB and red indicates high RoB.

### Appendix Table 3. Assessment of risk of bias for focus group

| Reference | Q1 | Q2 | Q3 | Q4 | Q5 | Q6 | Q7 | Q8 | Q9 | Q10 |
|---|---|---|---|---|---|---|---|---|---|---|
| Hinrichs, 2018[31] | | | | | | | | | | |

**Risk of bias criteria for focus group studies:** If any two of the items (i.e., research design, recruitment strategy, relationship between researcher and participants, data collection, ethical issues, data analysis, clarity of findings) had high RoB, the study was deemed high RoB. If a study had only one high risk item, it was deemed moderate RoB, and if it had no high-risk item, it was considered low RoB. **Questions:** Q1. Was there a clear statement of the aims of the research? Q2. Is a qualitative methodology appropriate? Q3. Was the research design appropriate to address the aims of the research? Q4. Was the recruitment strategy appropriate to the aims of the research? Q5. Was the data collected in a way that addressed the research issue? Q6. Has the relationship between researcher and participants been adequately considered? Q7. Have ethical issues been taken into consideration? Q8. Was the data analysis sufficiently rigorous? Q9. Is there a clear statement of findings? Q10. How valuable is the research? Green indicates an item as having low RoB and yellow indicates moderate RoB.

### Appendix Table 4. Percentage of CROSS reporting criteria satisfied by individual studies

| Section | Heading | Reporting criteria | Reporting criterion satisfied by studies (%) |
|---|---|---|---|
| Title and abstract | Title | 1a. State the word "survey" along with a commonly used term in title or abstract. | Clearly reported (58%)<br>Not reported (42%) |
| | Abstract | 1b. Provide an informative summary in the abstract. | Partially reported (100%) |
| Introduction | Background | 2. Provide a background about the rationale of study. | Clearly reported (100%) |
| | Purpose/aim | 3. Identify specific purposes, aims, goals, or objectives of the study. | Clearly reported (100%) |
| Methods | Study design | 4. Specify the study design in the methods section. | Clearly reported (75%)<br>Not reported (25%) |
| | Data collection methods | 5a. Describe the questionnaire. | Clearly reported (42%)<br>Partially reported (42%)<br>Unclearly reported (8%)<br>Not reported (8%) |
| | | 5b. Describe all questionnaire instruments that were used in the survey. | Clearly reported (25%)<br>Partially reported (75%) |
| | | 5c. Provide information on pretesting of the questionnaire, if performed. | Unclearly reported (17%)<br>Not reported (83%) |
| | | 5d. Questionnaire, if possible, should be fully provided. | Clearly reported (25%)<br>Partially reported (50%)<br>Unclearly reported (25%) |
| | Sample characteristics | 6a. Describe the study population. | Clearly reported (17%)<br>Partially reported (83%) |
| | | 6b. Describe the sampling techniques used. | Clearly reported (33%)<br>Partially reported (33%)<br>Not reported (34%) |
| | | 6c. Provide information on sample size, along with details of sample size calculation. | Clearly reported (34%)<br>Partially reported (58%)<br>Not reported (8%) |
| | | 6d. Describe how representative the sample is of the study population. | Clearly reported (34%)<br>Unclearly reported (8%)<br>Not reported (58%) |
| | Survey administration | 7a. Provide information on modes of questionnaire administration. | Clearly reported (75%)<br>Partially reported (17%)<br>Not reported (8%) |
| | | 7b. Provide information of survey's time frame. | Clearly reported (66%)<br>Not reported (34%) |

| | | | |
|---|---|---|---|
| | | 7c. Provide information on the entry process:<br>> For non-web-based surveys, provide approaches to minimize human error in data entry.<br>> For web-based surveys, provide approaches to prevent “multiple participation” of participants. | Not reported (100%) |
| | Study preparation | 8. Describe any preparation process before conducting the survey. | Clearly reported (8%)<br>Not reported (92%) |
| | Ethical consideration | 9a. Provide information on ethical approval for the survey if obtained. | Clearly reported (58%)<br>Partially reported (8%)<br>Not reported (34%) |
| | | 9b. Provide information about survey anonymity, confidentiality, and mechanisms used to protect unauthorized access. | Clearly reported (16%)<br>Partially reported (42%)<br>Not reported (42%) |
| | Statistical analysis | 10a. Describe statistical methods and analytical approach. | Clearly reported (67%)<br>Partially reported (25%)<br>Not reported (8%) |
| | | 10b. Report any modification of variables used in the analysis (if available). | Not reported (100%) |
| | | 10c. Report details about how missing data was handled. | Not reported (100%) |
| | | 10d. State how non-response error was addressed. | Clearly reported (8%)<br>Partially reported (8%)<br>Not reported (84%) |
| | | 10e. State how loss to follow-up was addressed for longitudinal surveys. | Not applicable since all studies were cross-sectional surveys. |
| | | 10f. Indicate whether any methods have been used to adjust for non-representativeness of the sample. | Clearly reported (8%)<br>Not reported (92%) |
| | | 10g. Describe any sensitivity analysis conducted. | Clearly reported (25%)<br>Unclearly reported (8%)<br>Not reported (67%) |
| Results | Respondent characteristic s | 11a. Report numbers of individuals at each stage of the study. | Clearly reported (92%)<br>Partially reported (8%) |
| | | 11b. Provide reasons for non-participation at each stage, if possible. | Clearly reported (34%)<br>Partially reported (8%)<br>Not reported (58%) |
| | | 11c. Report response rate, present the definition of response rate or the formula used to calculate response rate. | Clearly reported (8%)<br>Partially reported (84%)<br>Not reported (8%) |

| | | | |
|---|---|---|---|
| | | 11d. Provide definition and number of unique visitors. | Unclearly reported (8%)<br>Not reported (92%) |
| | Descriptive results | 12. Provide characteristics of study participants, and information on potential confounders and assessed outcomes. | Partially reported (92%)<br>Not reported (8%) |
| | Main findings | 13a. Give unadjusted estimates and, if applicable, confounder-adjusted estimates along with 95% confidence intervals and p-values. | Not reported (100%) |
| | | 13b. For multivariable analysis, provide information on the model building process, model fit statistics, and model assumptions (as appropriate). | Clearly reported (8%)<br>Not reported (92%) |
| | | 13c. Provide details about any sensitivity analysis performed. | Clearly reported (25%)<br>Not reported (75%) |
| Discussion | Limitations | 14. Discuss the limitations of the study, considering sources of potential biases and imprecisions. | Clearly reported (66%)<br>Partially reported (17%)<br>Not reported (17%) |
| | Interpretations | 15. Give a cautious overall interpretation of results and suggest areas for future research. | Clearly reported (8%)<br>Partially reported (92%) |
| | Generalizability | 16. Discuss the external validity of the results. | Clearly reported (34%)<br>Partially reported (33%)<br>Not reported (33%) |
| Other sections | Role of funding source | 17. State whether any funding organization has had any roles in the survey's design, implementation, and analysis. | Clearly reported (25%)<br>Not reported (75%) |
| | Conflict of interest | 18. Declare any potential conflict of interest. | Clearly reported (42%)<br>Not reported (58%) |
| | Acknowledgements | 19. Provide names of organizations/persons that are acknowledged along with their contribution to the research. | Clearly reported (50%)<br>Not reported (50%) |

### Appendix Table 5. Percentage of COREQ reporting criteria satisfied by studies

| Section | Heading | Reporting criteria | Reporting criterion satisfied by studies (%) |
|---|---|---|---|
| Research team and reflexivity | Personal Characteristics | 1. Interviewer/facilitator: Which author/s conducted the interview or focus group? | Clearly reported |
| | | 2. Credentials: What were the researcher's credentials (e.g., PhD, MD)? | Clearly reported |
| | | 3. Occupation: What was their occupation at the time of the study? | Clearly reported |
| | | 4. Gender: Was the researcher male or female? | Clearly reported |
| | | 5. Experience and training: What experience or training did the researcher have? | Not reported |
| | Relationship with participants | 6. Relationship established: Was a relationship established prior to study commencement? | Not reported |
| | | 7. Participant knowledge of the interviewer: What did the participants know about the researcher (e.g., personal goals, reasons for doing the research)? | Partially reported |
| | | 8. Interviewer characteristics: What characteristics were reported about the interviewer/facilitator (e.g., Bias, assumptions, reasons and interests in the research topic)? | Clearly reported |
| Study design | Theoretical framework | 9. Methodological orientation and Theory: What methodological orientation was stated to underpin the study (e.g., grounded theory, discourse analysis, ethnography, phenomenology, content analysis)? | Clearly reported |
| | Participant selection | 10. Sampling: How were participants selected (e.g., purposive, convenience, consecutive, snowball)? | Clearly reported |
| | | 11. Method of approach: How were participants approached (e.g., face-to-face, telephone, mail, email)? | Clearly reported |
| | | 12. Sample size: How many participants were in the study? | Clearly reported |
| | | 13. Non-participation: How many people refused to participate or dropped out? Reasons? | Not reported |
| | Setting | 14. Setting of data collection: Where was the data collected (e.g., home, clinic, workplace)? | Clearly reported |
| | | 15. Presence of non-participants: Was anyone else present besides the participants and researchers? | Not reported |
| | | 16. Description of sample: What are the important characteristics of the sample (e.g., demographic data, date)? | Clearly reported |

| | | | |
|---|---|---|---|
| | Data collection | 17. Interview guide: Were questions, prompts, guides provided by the authors? Was it pilot tested? | Partially reported |
| | | 18. Repeat interviews: Were repeat interviews carried out? If yes, how many? | Not reported |
| | | 19. Audio/visual recording: Did the research use audio or visual recording to collect the data? | Clearly reported |
| | | 20. Field notes: Were field notes made during and/or after the interview or focus group? | Clearly reported |
| | | 21. Duration: What was the duration of the interviews or focus group? | Not reported |
| | | 22. Data saturation: Was data saturation discussed? | Not reported |
| | | 23. Transcript returned: Were transcripts returned to participants for comment and/or correction? | Not reported |
| Analysis and findings | Data analysis | 24. Number of data coders: How many data coders coded the data? | Not reported |
| | | 25. Description of the coding tree: Did authors provide a description of the coding tree? | Not reported |
| | Respondent characteristic s | 26. Derivation of themes: Were themes identified in advance or derived from the data? | Clearly reported |
| | | 27. Software: What software, if applicable, was used to manage the data? | Not reported |
| | | 28. Participant checking: Did participants provide feedback on the findings? | Not reported |
| | Reporting | 29. Quotations presented: Were participant quotations presented to illustrate the themes / findings? Was each quotation identified (e.g., participant number)? | Clearly reported |
| | Main findings | 30. Data and findings consistent: Was there consistency between the data presented and the findings? | Clearly reported |
| | | 31. Clarity of major themes: Were major themes clearly presented in the findings? | Clearly reported |
| | | 32. Clarity of minor themes: Is there a description of diverse cases or discussion of minor themes? | Clearly reported |
